# Real-time optical analysis of a colorimetric LAMP assay for SARS-CoV-2 in saliva with a handheld instrument improves accuracy compared to endpoint assessment

**DOI:** 10.1101/2021.01.13.21249412

**Authors:** Lena M. Diaz, Brandon E. Johnson, Daniel M. Jenkins

## Abstract

Controlling the course of the COVID-19 pandemic will require widespread deployment of consistent and accurate diagnostic testing of the novel coronavirus SARS-CoV-2. Ideally, tests should detect a minimum viral load, be minimally invasive, and provide a rapid and simple readout. Current FDA-approved RT-qPCR-based standard diagnostic approaches require invasive nasopharyngeal swabs and involve laboratory-based analyses that can delay results. Recently, a loop mediated isothermal nucleic acid amplification (LAMP) test that utilizes colorimetric readout received FDA approval. This approach utilizes a pH indicator dye to detect drop in pH from nucleotide hydrolysis during nucleic acid amplification. This method has only been approved for use with RNA extracted from clinical specimens collected via nasopharyngeal swabs. In this study, we developed a quantitative LAMP-based strategy to detect SARS-CoV-2 RNA in saliva. Our detection system distinguished positive from negative sample types using a handheld instrument that monitors optical changes throughout the LAMP reaction. We used this system in a streamlined LAMP testing protocol that could be completed in less than two hours to directly detect inactivated SARS-CoV-2 in minimally processed saliva that bypassed RNA extraction, with a limit of detection (LOD) of 50 genomes/reaction. The quantitative method correctly detected virus in 100% of contrived clinical samples spiked with inactivated SARS-CoV-2 at either 1X (50 genomes/reaction) or 2X (100 genomes/reaction) of the LOD. Importantly the quantitative method was based on dynamic optical changes during the reaction so was able to correctly classify samples that were misclassified by endpoint observation of color.

## Introduction

Widespread deployment of rapid, accurate diagnostics is one of several key requirements for blunting the COVID-19 pandemic. Ideal tests are minimally invasive, requiring little or no sample preparation steps, and can be conducted on-site with a rapid and simple readout of a test outcome. However, the standard diagnostic approach requires an RNA extraction step from a nasopharyngeal (NP) swab and subsequent Reverse Transcription-quantitative Polymerase Chain Reaction (RT-qPCR). The invasiveness of the collection procedure often causes patient discomfort, reducing enthusiasm for regular retesting. Swab samples must then be stored in a viral transport medium (VTM) to stabilize the virus until the sample can be analyzed, usually after shipping to off-site laboratories. Furthermore, RNA extraction and RT-PCR analysis require specialized equipment and clinical laboratory training that are not easily adaptable to near-patient analysis. Finally, slow sample processing and nucleic acid amplification procedures cannot be significantly streamlined to facilitate rapid detection.

Testing saliva for RNA of respiratory viruses like SARS-CoV-2 has been shown to have high agreement in performance and sensitivity to approved RT-qPCR based tests of NP swab-based molecular tests.^1–3^ Isothermal analogs to PCR such as loop-mediated isothermal amplification (LAMP) can be implemented with comparatively simple and low-power equipment ^4–6^ and has proven to reliably amplify synthetic SARS-CoV-2 genomic RNA from samples with as few as five copies of template molecules per reaction.^7^ LAMP specifically is commonly observed to be more robust than equivalent qPCR assays to inhibitors in clinical samples, so that accurate results can be obtained even with rudimentary sample preparation (i.e. just add sample mixed with extraction buffer).^8–10^ Even so, as of December, 2020, less than 3% of all commercially available diagnostics with regulatory approval used for SARS-CoV-2 detection in the Unites States FDA, European Union and Asia utilize isothermal technology.^11^

The colorimetric SARS-CoV-2 LAMP diagnostic assay developed by COLOR was one of the first of these isothermal assays to receive Emergency Use Authorization (EUA) by the FDA.^12^ In this assay pyrophosphate hydrolyzed during a positive amplification event lowers the pH of the reaction buffer containing phenol red indicator, resulting in an observable change in color.^13^ This assay has proven to have a low false negative rate when used in conjunction with purified gamma-irradiated virus and purified viral clinical samples.^12^ While other FDA-EUA approved isothermal based methods have received FDA approval when used for serum or respiratory samples (i.e. nasopharyngeal swabs),^14–16^ approvals for direct detection in saliva have lagged behind no matter the molecular basis of the test. While SARS-CoV-2 RNA is as stable in saliva as it is in VTM,^1,17^ the effects of saliva on direct LAMP detection is poorly understood and VTM can interfere with the colorimetric readout.^18^ For example, variations in pH and buffering capacity of individual saliva samples can confound results of pH-dependent assays such as colorimetric LAMP, especially if relying on visual classification of endpoint color.

To date, the use of a colorimetric LAMP assay with clinical saliva samples has not been reported. Applications of isothermal amplification to detect SARS-CoV-2 have utilized commercially available polymerase enzyme mixes that contain nucleotides, Tris pH buffer, and Tween detergent. Additional studies have optimized amplification efficiency of genomic RNA from whole-virus samples resuspended in water by supplementing commercial reaction mixes with guanidinium chloride,^19^ Tris-EDTA (TE) and Tween-20.^20^ Detection sensitivity of whole virus has been further optimized by pre-incubating viral samples at 95°C to denature the viral capsid before adding samples to a LAMP reaction mix.^21–23^ Sample processing protocols have not been optimized for the direct detection of SARS-CoV-2 from saliva. In this work we report a streamlined approach for direct detection of SARS-CoV-2 RNA in contrived saliva samples, including simple sample preparation steps and monitoring of reactions in a hand-held instrument for dynamic detection of color changes.

## Materials and Methods

### SARS-CoV-2 RNA standards and controls

For the experiments in this study, contrived samples consisting of water, buffers or saliva were spiked with either synthetic RNA (BEI Resources catalog no. NR-52358, Lot no. 70035241) or gamma-irradiated (BEI Resources catalog no. NR-52287, Lot no. 70033322) SARS-CoV-2 virus. The reported starting genome copy number, designated throughout this study as genome equivalents (ge), for the synthetic RNA and gamma-irradiated SARS-CoV-2 are 1.05 × 10^8^ ge/mL and 1.7 × 10^9^ ge/mL (pre-inactivation) respectively. The following reagent was deposited by the Center for Disease Control and Prevention and obtained through BEI Resources, NIAID, NIH: Quantitative Synthetic RNA from SARS-Related Coronavirus 2, NR- 52358, SARS-Related Coronavirus 2, Isolate USA-WA1/2020 and Gamma-Irradiated, NR- 52287. The synthetic RNA includes fragments from the ORF 1ab, Envelope (E) and Nucleocapsid (N) regions. All virus stocks were aliquoted into 10 single use individual stocks and stored at -80°C. Stocks were diluted to the appropriate concentration in RNase/DNase-free water each day before experimentation.

### Quantitative analysis of colorimetric-luminance readout

#### Instrumentation

For real-time monitoring and quantitative detection of colorimetric LAMP assays we customized a handheld, battery-powered, fluorescence-based instrument with a heating block accommodating a strip of 8 PCR tubes (BioRanger™, Diagenetix Inc., Honolulu, HI, USA; Fig. 1A). The instrument interfaces to a companion Android app to monitor changes in color of the reaction mix. The modifications made to the device were removal of emission filters, and attenuation of raw signal by a combination of placing a diffuser over the photodiode detectors and electrically lowering their transimpedance gains. The resulting luminance signal (L) in arbitrary units (a.u.) results from scattering and reflections of excitation light not absorbed by the sample. Changes in the luminance in the dynamically monitored system correspond primarily to changes in absorbance of light by the sample. For sample illumination we tested three different color LEDs (Luxeon Rebel Color Line, Lumileds Ltd., San Jose, CA): blue (part number LXML- PB01-0040); green (part number LXML-PM01-0100), and; amber (part number LXML-PL01- 0040). Preliminary experiments (Supplementary Fig. S0) indicated that the green LEDs resulted in features in positive amplification curves that were identifiable earlier in the reactions than curves from other colors, and which could also be used for more definitive reaction classification. Based on these preliminary results we used green channel for monitoring reactions reported in this study, using quantitative analysis of amplification curves as described below. Calibration of the instrument was made with representative endpoint reactions with negative standards (pink solution, nominally assigned 0 a.u) and positive standards (yellow solution, nominally assigned 60000 a.u.).

**Figure 1.**
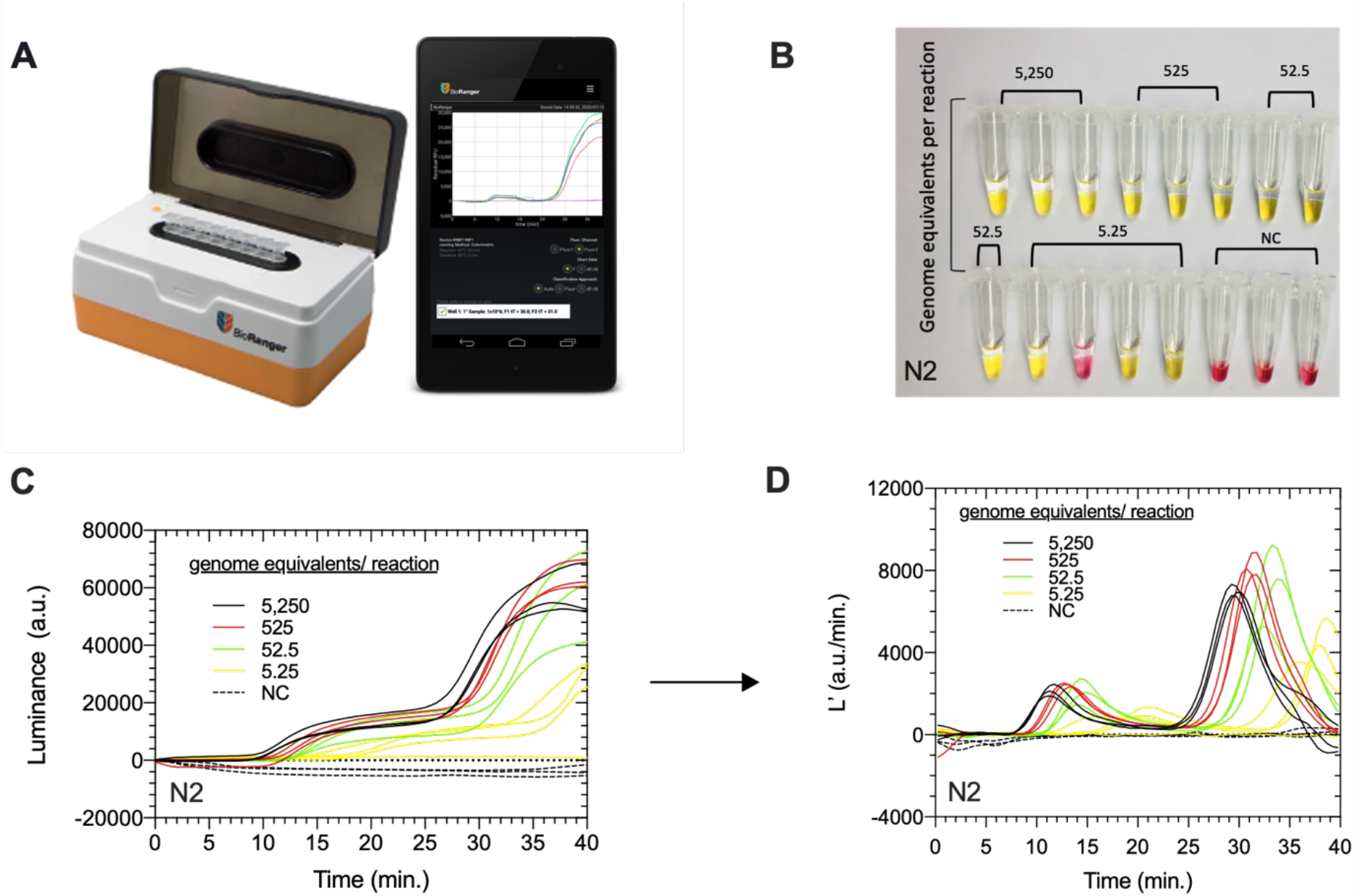
Visual and quantitative results of colorimetric-LAMP assay controls. **A**) BioRanger diagnostic platform modified for colorimetric LAMP, interfaced to a smartphone app that records luminance. **B**) Photograph of completed colorimetric LAMP assays performed under optimized reactions conditions for N-gene of synthetic SARS-CoV-2 RNA in water standards and corresponding **C**) luminance (L) and **D**) luminance derivative (L’) amplification curves as a function of time for each standard replicate.

#### Processing Luminance Data

For detailed analysis of experimental reactions described in this manuscript we used the raw “luminance” data from files generated automatically in the BioRanger app. Unless otherwise stated, all values were shifted to start at “zero” luminance by subtracting the initial (t = 0 minutes) raw luminance value from every raw value. These baseline corrected luminance values (L) were smoothed using a 2^nd^ order polynomial with a rolling average of 4 adjacent data points. The maximum values of luminance derivatives (L’) for each assay were identified from derivative data (identified using forward difference and smoothed again with a 2^nd^ order polynomial with 4 neighbors) using GraphPad Prism 9 (GraphPad Software, San Diego, CA, USA).

### LAMP assays

#### Primers

Two previously published LAMP primer sets (Supplementary Table S1) developed by New England BioLabs,^24^ targeting the envelope (E1) and nucleocapsid (N2) genes of the SAR- CoV-2 viral genome (GenBank accession number MN908947), were tested individually and in combination. For the final contrived clinical evaluation an internal control (IC) primer set that amplifies human β*−*actin “housekeeping” gene (ACTB) was used to detect the presence of inhibitors in saliva samples. Primers were synthesized commercially (Integrated DNA Technologies / IDT, Coralville, Iowa) using standard desalting and resuspended in nuclease free water. For each primer set an individual 10X primer stock was prepared so that adding 2.5 *µ*L of each stock to a LAMP reaction yielded the following final primer concentrations in both fluorescent and colorimetric LAMP assays: 0.2 *µ*M F3/B3, 1.6 *µ*M FIP/BIP and 0.8 *µ*M LB/LF. For LAMP assays containing dual primer sets targeting both the E and N gene of SARS-CoV-2 genome, 2.5 *µ*L of each 10X stock were added to each reaction mixture.

#### Colorimetric LAMP Assay

All colorimetric assays contained 12.5 *µ*L WarmStart^®^ RT-Colorimetric 2X Master Mix (RNA & DNA, M1800, New England Biolabs, MA, USA), 2.5 *µ*L primer mix (10X) stock of each primer set used (E1, N2 or ACTB), either 2 *µ*L or 5 *µ*L of sample and appropriate volumes of nuclease free water to make a total volume of 25 *µ*L per reaction. After loading LAMP reagents and test samples a drop (∼ 20 *µ*L) of sterile mineral oil was added to each reaction tube (excluding some of the negative controls in preliminary evaluation) and closed, followed by brief centrifugation of the 8-assay tube strip.

For reactions containing guanidine hydrochloride (GuHCl molecular grade, J7582322, ThermoFischer Scientific, Waltham, MA, USA), 1.25 *µ*L of GuHCl stock (0.8 M) was substituted for the equivalent volume of nuclease free water to achieve a final reaction concentration of 40 mM GuHCl. Stock GuHCl solution was prepared in deionized water and adjusted to pH 8.0 with 1 M KOH (90% reagent grade, 484016, Sigma Aldrich, St. Louis, MO, USA) and filter sterilized (0.22 *µ*M 13 mm Whatman filter, 99091302, MilliporeSigma, St. Louis, MO, USA).

All real-time colorimetric assays were carried out in 0.2 mL reaction tubes (TempAssure, Optical Caps, USA Scientific Inc., Orlando, FL, USA) in a modified 8-well isothermal amplifier platform (BioRanger). Amplification progress was logged every 30 seconds for a maximum of 40 minutes. At the end of the assay the closed tube strip was briefly placed on ice and a photograph was taken using a cell phone camera. The colorimetric master mix contains a phenol red pH indicator used for visual detection of amplification and interpreted according to manufacturers (NEB) protocols^25^ as follows: negative reactions remain pink while successful amplification results in yellow or yellow/orange color observable by naked eye.

#### Fluorescent LAMP Assay

Fluorescent LAMP assays were performed in parallel to most colorimetric assays under the same experimental and assay conditions except fluorescent assays included 0.5 *µ*L fluorescent dye (50X, E1700S, New England Biolabs). Assays were performed in either a) 0.2 mL reaction tubes (TempAssure, Optical Caps, USA Scientific Inc., Orlando, FL, USA) in an *unmodified* 8-well isothermal amplifier platform (BioRanger™, Diagenetix Inc., Honolulu, HI, USA) or b) 0.1 mL reaction tubes (TempAssure, Optical Caps, USA Scientific Inc., Orlando, FL, USA) in a real-time PCR machine (Applied Biosciences StepOnePlus™). Both instruments were programmed for isothermal incubation at 65°C for 31 minutes. Fluorescence values corresponding to fluorescein were recorded every minute during the 31-minute reactions in the StepOnePlus™ PCR machine, and every 30 seconds in the BioRanger isothermal amplifier. For the BioRanger, threshold times (t_T_) of positively classified reactions are reported as the time at which the maximum rate of fluorescence increase occurs, to control for variations in reaction intensity and fluorescence sensitivity between channels. For the StepOnePlus™ PCR machine, the t_T_ was estimated as the time required for the fluorescence value to exceed a threshold value equivalent to the pooled average plus three standard deviations of the fluorescence values observed throughout the reactions of triplicate negative control reactions.^26,27^

### Quantitative colorimetric-LAMP assay development

#### Controls and Standards

For preliminary assay optimization and characterization, positive control (PC) standards containing 5.25 – 5,250 ge/ 5 *µ*L were prepared daily by dissolving synthetic SARS-CoV-2 RNA from frozen stock in nuclease free deionized water, whereas negative controls (NC) did not contain target template RNA. For contrived clinical sample testing reactive controls (RC) containing 5 × 10^3^ ge/mL – 1.0 × 10^6^ ge/mL in the primary sample were prepared by spiking saliva with inactivated (gamma-irradiated) SARS-CoV-2 RNA and non-reactive control (NRC) samples are saliva without added template.

#### Reaction Condition Optimization

Fluorescent LAMP assays performed on a StepOnePlus™ PCR machine was used to evaluate the effect of adding GuHCl, compare sensitivity of different LAMP primers, and determine optimal reaction temperature for detection of viral RNA in water standards. 5 *µ*L samples of each PC and NC standard was added to a LAMP assay to achieve 0 – 5,250 ge/reaction with or without GuHCl and incubated for 31 minutes at either 65°C or 68°C. Primer sets were evaluated individually and duplexed (E1 + N2) by adding equal volumes (1:1) of stock primer mixes to a LAMP reaction. The Limit of Detection (LOD) for each set of assay and processing conditions was determined as the lowest concentration where detection occurred in 3/3 replicates of PC reactions.

#### Quantitative Classification of Results from Luminance Data

To identify features in luminance curves that consistently differentiated positive and negative colorimetric LAMP reactions, we considered simple quantitative metrics from amplification data recorded on the modified BioRanger device assaying RNA standards in water. The value of each metric for discriminating the binary populations was evaluated in a commercial software (GraphPad Software, San Diego, CA, USA) using unpaired t-test assuming equal variances.

Based on these preliminary results, Reactions were classified based on the value of the peak luminance derivative L’ (a.u./min.), as strong peaks are consistent with sigmoidal amplification characteristic of qPCR or LAMP. Classification thresholds were empirically derived from baseline measurements of controls standards without viral RNA. The threshold for positive classification was determined as the value excluding at least 99.9% of negative controls (for standards in water prepared with viral RNA) or non-reactive controls (for contrived clinical samples in saliva) based on observed means and sample standard deviations using a one-tailed t-statistic. The LOD for colorimetric assay in standards or contrived clinical samples in the modified instrument was defined as the lowest concentration (ge/mL) where all replicates were positively detected by this metric.

### Application of the quantitative colorimetric-LAMP assay for SARS-CoV-2 in saliva

#### Saliva Collection

Approximately 1-3 mL of saliva was collected from healthy individual volunteers in 15 mL conical tubes (352196, Falcon™, Thermo Fisher Scientific, Waltham, MA, USA). Donors were asked to not eat or drink anything other than water before saliva was collected. Saliva samples were collected fresh on the mornings of experimentation and stored at 4 °C until processed. On the day of each experiment saliva samples were spiked with known quantities of inactivated (gamma-irradiated) SARS-CoV-2 virus following protocols described in this section.

#### Standardized Processing and Assay of Contrived Clinical Samples

In the absence of known positive COVID-19 clinical samples, we followed the Federal Food and Drug Administration’s (FDA) guidance for molecular diagnostic development for Emergency Use Authorization^28^ and validated the sensitivity and accuracy of the colorimetric LAMP assay using contrived clinical samples. Saliva collected from three healthy donors presumed to be free of SARS-CoV-2 virus was pooled and spiked with inactivated gamma irradiated SARS-CoV-2. Spiked saliva samples were heat inactivated in a dry bath at 95 °C for 30 minutes, allowed to cool to room temperature then stored on ice until downstream processing. 100 *µ*L of each treated 1 mL spiked sample was combined with an equal volume (1:1) of stabilization buffer containing 1X TE (10 mM Tris, 0.1 mM EDTA, pH 8, IDT) and 1% Tween- 20 for a final sample concentration of 0.5X TE and 0.5% Tween-20. 2 *µ*L of each sample was added directly to the colorimetric LAMP reaction mix for detection of the SARS-CoV-2 N-gene using the N2 primer set only. Reactions were analyzed by real-time colorimetric LAMP (65°C, 40 minutes) on a BioRanger diagnostic platform modified to measure luminance of colorimetric reactions.

### Validation in contrived clinical samples

Ten individual saliva specimens were collected from healthy donors and evaluated for the real-time quantitative SARS-CoV-2 colorimetric LAMP assay. One milliliter of each of the 10 specimens was spiked with gamma-irradiated SARS-CoV-2 to produce five contrived positive clinical samples with a viral load at 1X the previously established LOD of 5 × 10^4^ ge/mL corresponding to 50 ge/reaction and five contrived positive samples at 2X the LOD (1 × 10^5^ ge/mL) corresponding to 100 ge/reaction. Unadulterated volumes from each original specimen were assayed identically to serve as a paired non-reactive sample for each contrived positive clinical sample. The 20 samples (10 negative and 10 positive) were randomized and blinded and processed through the entire standardized assay procedure.

For every saliva sample (n=20) an eight-assay diagnostic test panel was performed to simultaneously test two replicates each of the test sample, internal control (IC), positive control (PC) and negative control (NC). Saliva samples were tested for SARS-CoV-2 N-gene and human β-actin (IC). Positive controls contained 2.1 × 10^6^ ge/reaction of synthetic SARS-CoV-2 RNA (BEI 52358) dissolved in deionized water, and deionized water no template negative controls were both tested for the N-gene. Each reaction in the panel contained 2 *µ*L of sample, 40 mM of GuHCl and standard colorimetric master mix and primer concentrations as previously described. Assays were carried out and interpreted blind to the experimenter. The corresponding viral load in each sample for analysis was revealed at the end of the experiment only after the samples had been analyzed by quantitative colorimetric LAMP. The results of each diagnostic panel were interpreted by the experimenter both visually following the manufacturers protocol and quantitatively based on luminance signals for each individual LAMP assay.

## Results

### Reaction conditions

Fluorescent LAMP assays were used to evaluate optimal reaction conditions for assays containing single and duplexed primer sets. The addition of 40 mM GuHCl to fluorescent LAMP assays improved both the speed and sensitivity for the detection of synthetic RNA in spiked water controls. Assays containing only the N2 primer set and guanidine hydrochloride performed best overall in terms of sensitivity (∼ 5.25 genome equivalents/reaction) and speed of detection (mean t_T_ = 19 minutes, Supplementary Fig. 1). The differences in the speed of detection for duplexed reactions were unremarkable; E1 primers are less sensitive than the N2 primers by approximately an order of magnitude in any condition. E1-primed reactions were also more temperature sensitive: incubation at 68°C inhibited LAMP reactions. Colorimetric assays were run ten minutes longer than fluorescent assays for a total time of 40 minutes to compensate for the relatively slow color change of the phenol red indicator that has been demonstrated to take forty to fifty minutes to detect single copy numbers of SARS-CoV-2 genomes in previous studies.^29,30^ All subsequent colorimetric assays were performed under optimal conditions: 40 mM GuHCl and incubated at 65°C for 40 minutes.

### Visual and quantitative evaluation of assay controls

Colorimetric LAMP assays using the E1 primer set had relatively poor sensitivity and slow amplification (Supplementary Fig. 1, 2A-C). In comparison assays duplexed with both E1 and N2 primer sets were more sensitive but resulted in relatively noisy luminance profiles especially during the first five minutes of the reaction (Supplementary Fig. 2D-F). By comparison assays with just the N2 primer set resulted in smooth luminance profiles and detection of 5.25 ge/reaction in two out of three replicates (Fig. 1B-C). Luminance derivative (L’) profiles for positive standards amplified with N2 primer set had two identifiable peaks in contrast to negative reactions which had relatively flat luminance profiles (Fig. 1D). We subsequently chose to use the N2 primer set only in standardized reactions because of the robust and clear luminance signals and sufficient performance to detect the vast majority of SARS-CoV-2 positive samples based on reported saliva viral loads averaging 10^2^-10^3^ copies per microliter.^1,31^

**Figure 2.**
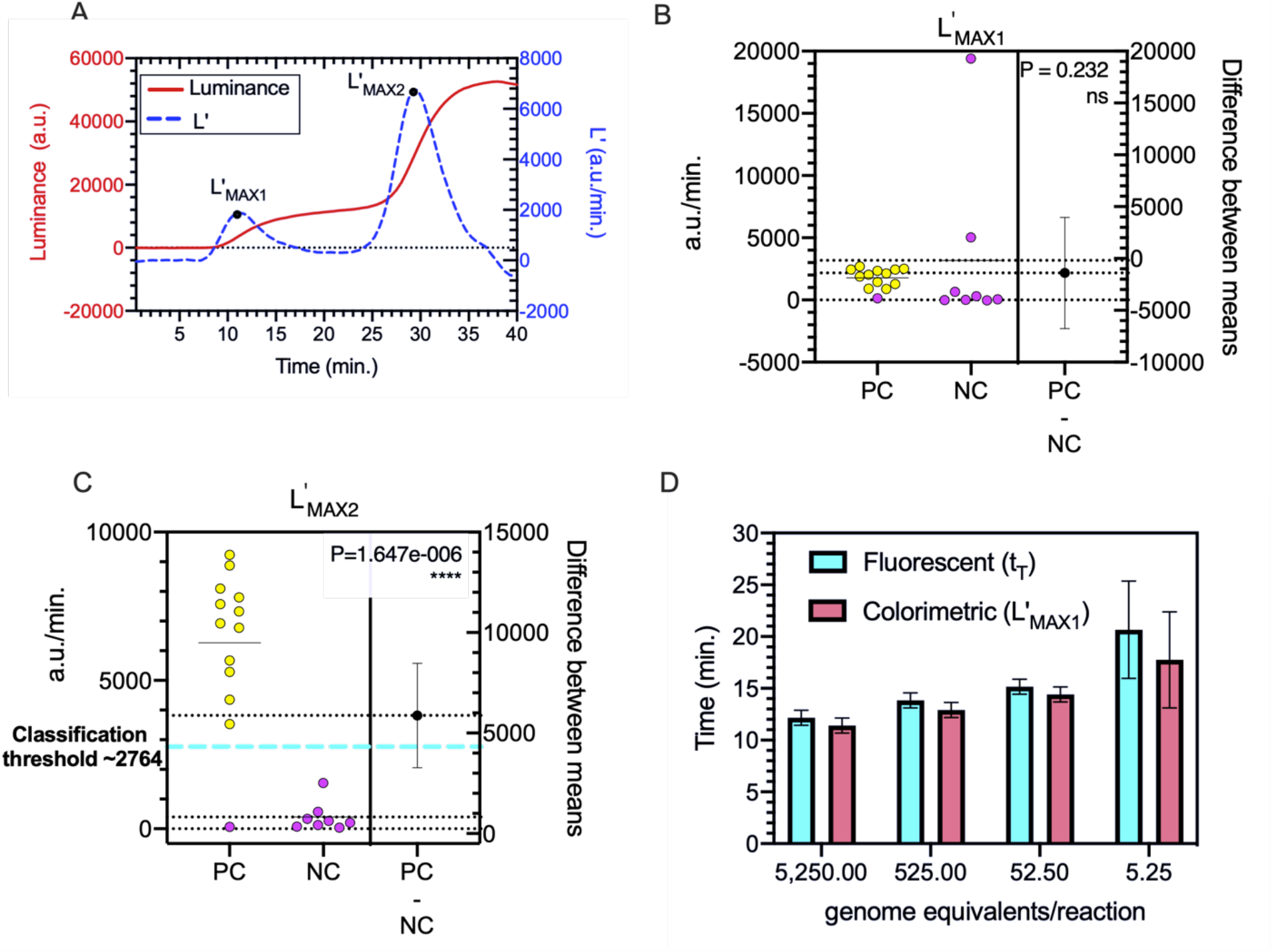
Quantitative analysis of positive (PC) and negative control (NC) synthetic RNA standards in water. **A)** Example luminance (L) curve (left y-axis) and corresponding derivative (L’, right y-axis) of an optimized positive control assay containing 5,250 genome equivalents and targeting the N-gene of synthetic SARS-CoV-2 RNA. The two derivative curve peaks indicate local maxima in rate of change in luminance, measured in arbitrary units (a.u.) between 5-20 minutes (L’_MAX1_) and between 20-40 minutes (L’_MAX2_). **B)** Scatter plot of first (L’_MAX1_) and **C)** second (L’_MAX2_) local maxima in luminance derivative (L’) values (a.u./min.). PC (n=13) and NC assays (n=8) were significantly (**** = P < 0.0005) different based on L’_MAX2._ The N-gene was quantitatively detected in 12/13 PC reactions containing a range of synthetic RNA (5.25 – 5,250 ge/reaction) in which L’_MAX2_ > ∼ 2764 a.u./min. (classification threshold = blue dash line). Symbols represent one assay replicate; color corresponds to endpoint reaction color and lines represent the mean. The difference between PC and NC (PC-NC, right y-axis) contains three values: the lower and upper 99% confidence level (dotted lines) and the difference between the means of the two groups (error bars) **D**) Threshold values (t_T_) corresponding to the time of detection of the N-gene in PC LAMP assays performed in parallel on a commercial fluorescence- enabled BioRanger correlated strongly to the time when L’_MAX1_ occurs. Error bars are standard error of mean (SEM).

Under optimized assay conditions the colorimetric luminance curve of PC standards is bi- phasic with two distinct exponential growth phases in the first and last half of the reaction. We quantitatively evaluated two curve features, presumably associated with initial amplification and subsequent color change, at time intervals where a log-linear luminance profile was observed to occur in PC assays. Specifically, local maxima in rates of luminance increase (L’) occur between 5 and 20 minutes (L’_MAX1_) and then again between 20 and 40 minutes (L’_MAX2_) (Fig. 2A). The first five minutes of luminance values were excluded from consideration for L’_MAX1_ because noise in the baseline signal was previously observed to occur early in the reaction, particularly for assays containing duplexed primer sets (Supplementary Fig. 2B, 2E).

The luminance curves of NC assays testing N2 primers (Fig. 1) plus five NC assays from other trials conducted that day testing different primers sets (Supplementary Fig. 2) were pooled (n=8) and compared to PC purified RNA standards (n=13, Fig. 1C-D). L’_MAX2_ was the only L’- maxima that significantly (P = 1.647e-006) differentiated PC from NC based on differences between means (PC - NC = 5,876 a.u./min., Fig. 2B, 2C). Correspondingly our decision threshold for classifying positive control RNA standards was L’_MAX2_ > 2764 a.u./min., the approximate equivalent to the pooled average of eight negative control assays (μ=394.4 a.u./min.) plus 4.78 sample standard deviations (sd=495.7, t_.999_=4.78, dof=7, α =0.001). Using this threshold, results from the colorimetric instrument are in 100% agreement with endpoint visual assessment of yellow color, with both approaches detecting 3/4 assays containing 5.25 ge/reaction with the N2 primer set, and every positive standard with more template RNA.

The times corresponding to the first luminance derivative peak (L’_MAX1_) in the colorimetric assay (10 – 20 minutes) were similar to threshold times observed in fluorescence-based LAMP assays with the same primers (Fig. 2D). These results contrast sharply with previous studies showing that incubation times of forty to fifty minutes are required for reliable color development and end-point detection of SARS-CoV-2 colorimetric LAMP assays.^12,29,32^ This indicates that our simple instrument is able to detect subtle changes in the optical characteristics of the reaction that occur before easily observable color changes. The second luminance derivative peaks (L’_MAX2_) in our instrument are much larger than L’_MAX1_ and are typically observed between 25-35 minutes into the reaction which is more consistent with times required for endpoint assays, suggesting that this shift is primarily due to the spectral shift in the pH indicator.

### Limit of detection in saliva

The sensitivity of the colorimetric-LAMP assay was quantitatively assessed by following optimized assay protocols for detection of SARS-CoV-2 in saliva using a handheld instrument for luminance readout (Fig. 3). To reduce the impact of saliva-bound nuclease activity, we supplemented the reaction buffer with Tris-EDTA (TE) to stabilize the viral genomic RNA released from the capsid following heat treatment of viral samples. EDTA protects nucleic acids from degradation by chelating the nuclease cofactor Mg2+ and Tris maintains a pH above 7.5, at which nucleases are less active. Researchers at the University of Illinois extensively evaluated cheaper alternatives to commercial sample transport buffers containing expensive and proprietary viral RNA-stabilizing agents like DNA/RNA Shield (Zymo Research), and demonstrated that TE buffer is effective at stabilizing nucleic acids for sensitive detection (500- 1000 viral particles per mL) by RT-qPCR.^20^ We similarly demonstrated that addition of TE to heat-inactivated samples containing intact gamma-irradiated SARS-CoV-2 improved the LOD of the colorimetric assay by an order of magnitude compared to spiked samples diluted in water only (Supplementary Fig. 3). Furthermore, their research suggests that extended heating of the sample (i.e. 30 minutes, 95°C) inactivates inhibitors in saliva whereas standard protocols for heat inactivation of SARS-CoV-2 at lower temperatures (i.e. 30 minutes, 60°C) did not allow for sensitive detection. Herein spiked saliva samples are subjected to heat treatments at 95°C for 30 min prior to diluting the sample 1:1 with TE buffer.

**Figure 3.**
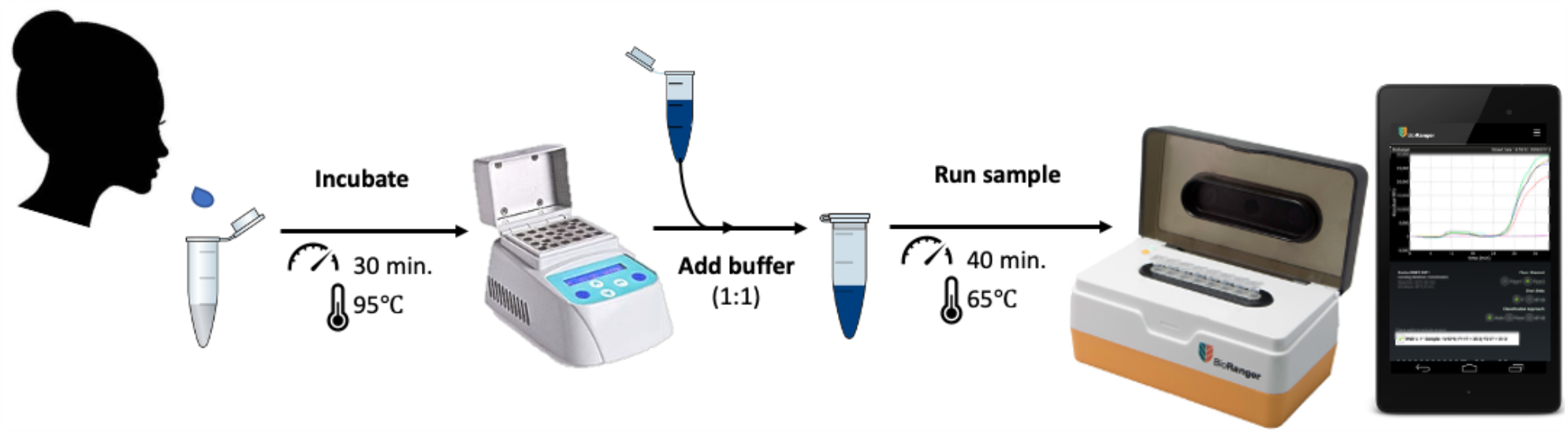
Schematic representation of colorimetric LAMP workflow to quantitatively detect SARS-CoV-2 in saliva using a handheld isothermal diagnostic platform. The optimized saliva testing protocol is as follows: 1-2 mL of saliva is collected, heat inactivated (95 °C for 30 minutes) then combined with equal parts TE buffer (1X, 1% Tween 20). Next 2 *µ*L of the diluted saliva sample is directly pipetted into a reaction tube containing Mastermix and primers and incubated (65 °C for 40 minutes) on the BioRanger: a mobile isothermal diagnostic platform capable of performing eight assays in parallel. Real-time luminance data is recorded on a custom smartphone app for post-test analysis of the results.

Reactive controls (RC) containing inactivated SARS-CoV-2 in saliva were classified as positive for detection if L’_MAX2_ was greater than 660 a.u./min, equivalent to the approximate average (μ=86.09 a.u./min.) plus 4.78 standard deviations (sd=119.1, t_.999_=4.78, dof=7, α =0.001) of eight non-reactive control (NRC) (Fig. 4A). Applying this classification threshold, the luminance-based LOD, defined by the lowest concentration where SARS-CoV-2 is detected in all replicates, is 5 × 10^4^ ge/mL corresponding to 50 ge/reaction (Fig. 4B). Luminance amplification curves for this section can be found in Supplementary Figure 4.

**Figure 4.**
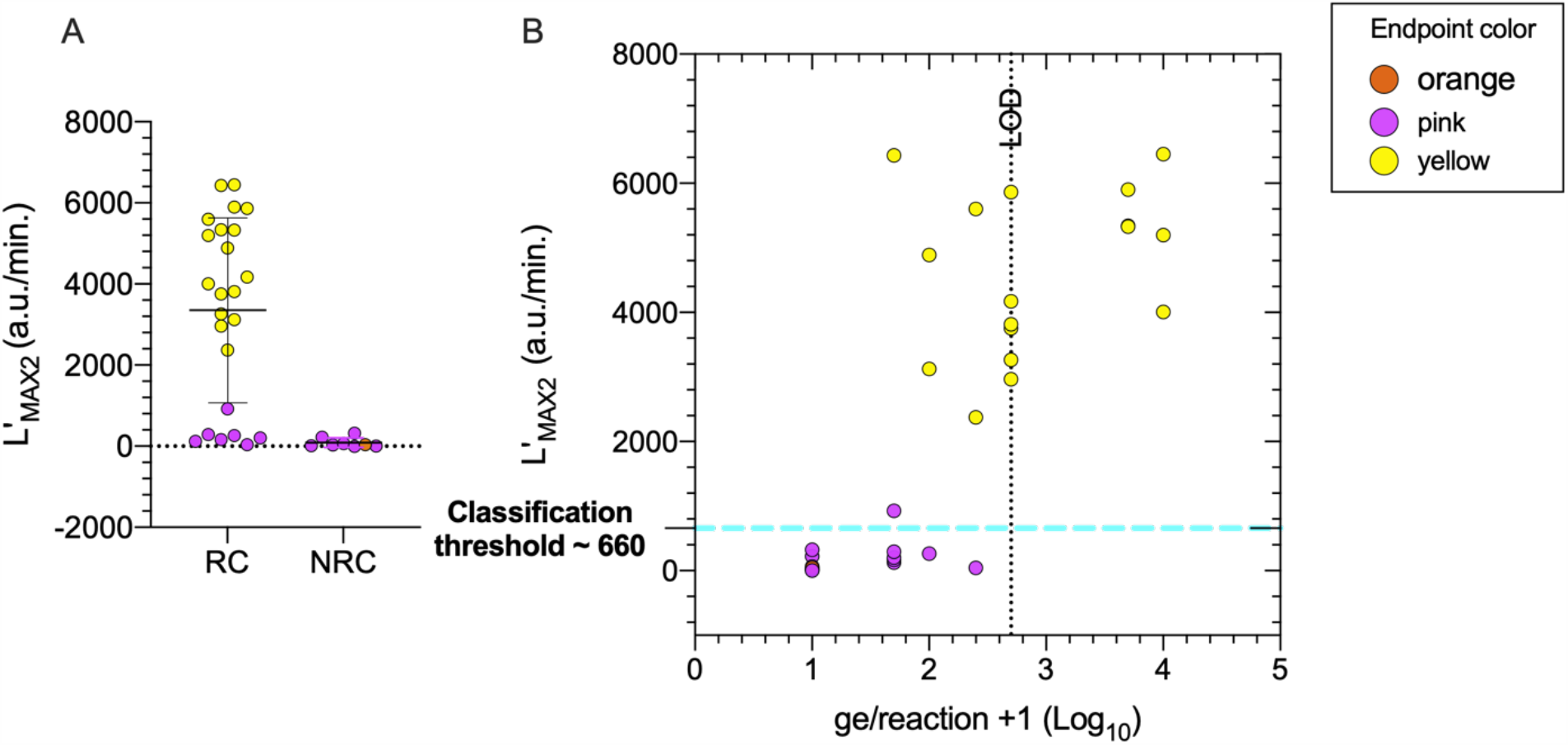
Quantitative and visual detection sensitivity of inactivated SARS-CoV-2 in saliva. **A)** Comparison of maximum amplification rates (L’_MAX2_) between reactive controls (RC) containing viral RNA and non-reactive control (NRC) saliva samples. Error bars represent mean and standard deviation. **B)** The LOD (dotted line; lowest concentration where the N-gene is detected in all replicates) by endpoint and luminance-based detection (L’_MAX2_ > 660 a.u./min.) is 50 ge/reaction equivalent to 5 ×10^4^ ge/mL in the primary sample. The color of each symbol, representing an assay replicate, corresponds to the endpoint color of the assay.

SARS-CoV-2 was detected both visually and quantitatively below the LOD (5 × 10^3^ ge/mL – 2.5 × 10^4^ ge/mL) in some replicates but at a lower rate. In some reactive samples below the LOD and in reactions in which SARS-CoV-2 was not visually detected, a low-amplitude luminance signal (L’_MAX2_ < 1,000 a.u./min) is observed in the last twenty minutes. This could indicate latent target amplification and a longer reaction time could improve sensitivity of small quantities of viral RNA. However longer incubation times (> 40 minutes) have shown to increase the false positive rate. One NRC reaction showed a color change, however the L’_MAX2_ values were less than 100 a.u./min. The yellow/orange NRC reaction was quantitatively classified as ‘Not Detected’ reducing the initial false positive (FP) rate of visual end-point determination from 12.5% (1/8 NRC turned orange/yellow) to 0%. The color change in the negative control may have occurred from the saliva destabilizing the buffer system demonstrating the importance of quantitative approaches to reduce the prevalence of false positives.

### Blind evaluation of the colorimetric LAMP assay using clinically contrived saliva samples

To evaluate the feasibility and performance of our quantitative colorimetric LAMP method, a blinded and randomized contrived clinical trial was conducted. A total of 20 saliva samples, 10 reactive controls containing inactivated SARS-CoV-2 at 1X and 2X the LOD (5 × 10^4^ ge/mL) determined from saliva standards (Fig. 4), and 10 non-reactive controls (0 ge/mL), were tested using the standardized protocol (Fig. 3). In addition to two replicate assays for the N-gene, each sample was subjected to a test panel including two replicates each of an internal control reaction (IC; for human β-actin), a positive control (PC; supplemented with 10^6^ ge/reaction synthetic RNA), and a negative control with no sample or supplemental RNA. Each individual assay of the test panel was interpreted both visually (pink = Not detected, yellow/orange = Detected) as well as quantitively, using the previously established luminance readout classification threshold determined from saliva standards (L’_MAX2_ > 660 a.u./min = Detected, L’_MAX2_ < 660 a.u./min. = Not detected). A test panel was interpreted as ‘Detect’ if SARS-CoV-2 N-gene was detected in 2/2 replicates, ‘Detected†’ for 1/2 replicates and ‘Non-detect’ for 0/2 replicates. Additionally, the panel results were deemed valid if both replicates for IC (2/2) and PC (2/2) were detected and neither NC (0/2) replicates were detected. Panels with any unexpected outcome of control reactions is classified as ‘Inconclusive’. Examples of three test panels are shown in Figure 5; detailed results for all 20 diagnostic panels are included in Supplementary Figure 5, and the sample pairing key is shown in Supplementary Table 2.

**Figure 5.**
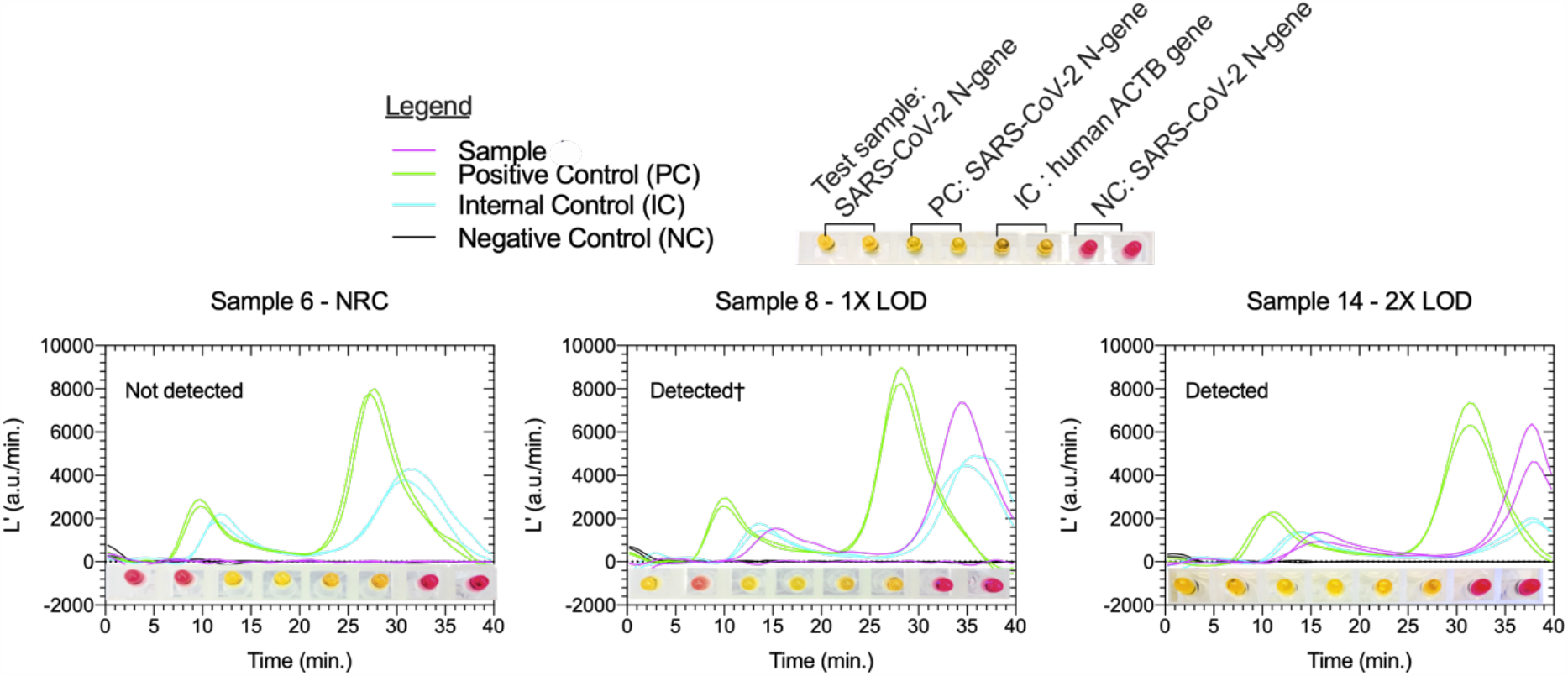
Example test panel configuration and results for each three sample types evaluated in the contrived clinical testing trial. Test panel results are interpreted both visually (pink = target not detected, yellow = target detected) and by luminance derivatives (L’). The test panels are interpreted based on the detection rate of SARS-CoV-2 N-gene in tested saliva samples as : ‘Not detected’ (0/2), ‘Detected†’ (1/2) or ‘Detected’ (2/2) if PC and IC assay targets are detected in all replicates and not detected in any NC replicates. In these three panels shown, visual and quantitative results are in 100% agreement for all assays. The results of all 20 saliva test panels can be found in Supplementary Figure 5.

Although the test panels were run on two colorimetry-enabled BioRanger instruments over two weeks and by multiple experimenters using different batches of reagent stocks, performance analysis demonstrates that the method sensitivities, specificities and accuracy to quantitatively detect SARS-CoV-2 did not vary substantially. All assays that resulted in an endpoint color of yellow also showed luminance signal amplification rate above decision threshold values where L’_MAX2_ was above 660 a.u./min. (Fig. 6A). A failed color change at the LOD correlated with no or low intensity luminance signals (L’_MAX2_ < 660). The SARS-CoV-2 N- gene was detected in all test sample replicates (10/10) for saliva samples spiked with 1 × 10^5^ ge/mL (2X LOD). At least one replicate tested positive for each sample with 5 × 10^4^ ge/mL (1X LOD), though not all replicates at this concentration tested positive (6/10) (Fig. 6B). SARS-CoV-2 was not detected in any of the contrived negative samples (NRC). Human ACTB gene was detected in all but one test panel IC (19/20 samples, 38/40 replicates).

**Figure 6.**
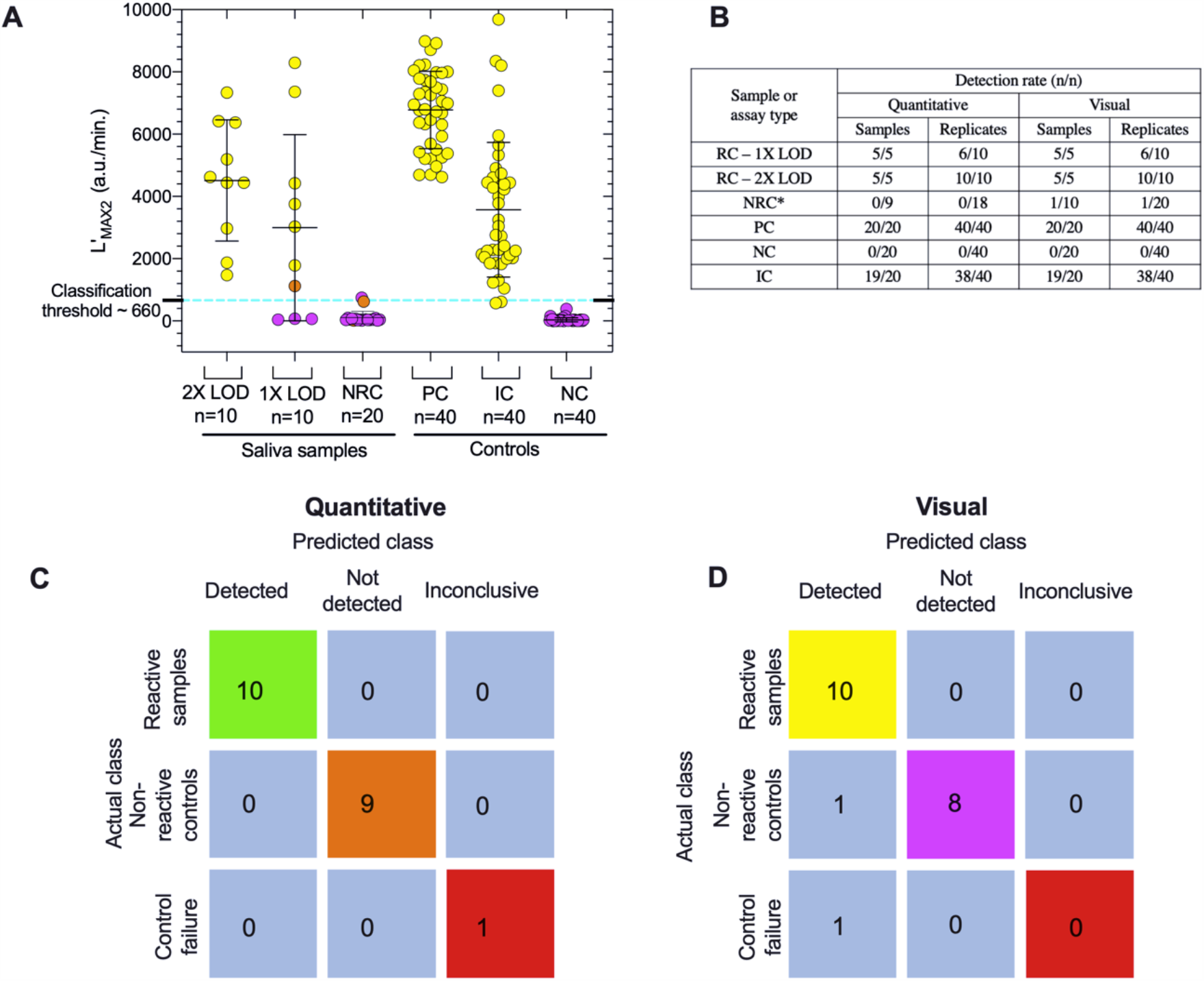
Performance analysis comparison of the quantitative colorimetric LAMP assay to visual detection using twenty clinical SARS-CoV-2 saliva test panels. (**A)** Distribution summary of L’_MAX2_ values for each tested saliva sample and control group used in the clinical evaluation trial. Blue dotted line represents classification threshold value (L’_MAX2_ > 660 a.u./min.) used to quantitatively determine whether the assay target is positively detected. Group means are indicated by lines, error bars are the standard deviation and the symbol color corresponds to the endpoint reaction color (orange, pink, yellow). (**B**) Table comparing visual to quantitative sensitivity in terms of sample and replicate detection rates for every sample and control group. ^*^NRC sample sizes are different due to IC failure only recognized by quantitative analysis of panel 9 (**C, D)** Confusion tables comparing quantitative and visual test panel interpretations (SARS-CoV-2 Detected, Not Detected or Inconclusive) made blindly by the experimenter to the actual classification of tested samples.

Sample 1 and sample 2 (Supplementary Fig. 5) were the first panels run on the day of testing and performed in parallel on two different instruments without first pre-heating the incubation chamber. This resulted in significant amount of noise in all assays of Panel 1 and one NRC assay of Panel 2. Panel 2 NRC assay was noisy from 0-25 minutes, resulting in an L’_MAX2_ value of 746 a.u./min. However, after inspection of the smoothed and corrected L’ curve, it was clear that the local maxima (L’_MAX2_) was an artifact due to noise and the assay was classified as ‘Not-detected’. Subsequently all tests were performed on pre-heated incubation blocks reaching a temperature of at least 50 °C prior to loading the reaction tubes. This phenomenon was observed in preliminary experiments but not well characterized and in most cases, the luminance curve corrections and smoothing were sufficient to remove noise or artifacts falsely indicating amplification.

Panel 9 tested an NRC sample, and even though endpoint assessment of the color (light orange) resulted in positive classification for both replicates, no signs of amplification were observed in the luminance data. Using the real-time data, both classified correctly as negative (Supplementary Fig. 5, panel 9). Similarly, one replicate of the panel 3 sample (also an NRC sample) was light orange incorrectly indicating amplification based on endpoint color, though the luminance data correctly showed no amplification (Supplementary Fig. 5, panel 3). In contrast the internal control reactions on panel 9 showed evidence of faint and late amplification in the luminance data but failed to meet threshold for positive classification, resulting in this panel being inconclusive, even though the endpoint color of these internal controls was yellow (positive). The N-gene was correctly detected in all positive control replicates (40/40) and not detected in any of the negative control replicates (0/40) (Fig. 6A-B).

The L’_MAX2_ values provide a robust indication of nucleic acid amplification that may not accompany an endpoint color change that is sensitive to variations in sample characteristics. This suggests that dynamically monitoring luminescence might improve classification accuracy where sample pH is too low, in addition to situations in which buffering capacity is too high to yield a complete red to yellow color change. In comparison, classification based on endpoint color in our panels results in a false positive rate of 10% near the LOD.

## Discussion

As COVID-19 continues to surge, there is increasing need to expand points-of-care and mobile testing platforms to quell the transmission of this virus. Nucleic acid amplification emerged as the gold standard for detection efficiency in the early months of the pandemic because this method is both highly specific and highly sensitive. Paired with an RNA isolation sample-processing step, RT-PCR-based SARS-CoV-2 detection assays provided the necessary data for viral transmission rates and initial contact tracing. RT-PCR-based testing, while accurate and sensitive, is heavily reliant on thermocycling equipment that requires a specialized laboratory environment and highly trained personnel to operate. These limitations have hindered effort to scale the use of this testing platform to meet the exponential expansion in testing demand. Antibody-based detection platforms are emerging as an alternative to RT-PCR based assays to meet the rising demand for rapid, point-of-care testing. While antibody-based detection methods can be adapted for at-home, point-of-care detection, or mobile testing, this methodology is inherently less sensitive than nucleic acid amplification-based assays.

Isothermal nucleic acid amplification merges the mobility of antibody-based assays with the high sensitivity of RT-PCR as an efficient platform to expand COVID-19 testing. In this study, we have built upon the colorimetric endpoint readout of LAMP-based testing and have provided proof-of-design for a mobile platform that utilizes isothermal nucleic acid amplification in conjunction with direct detection of SARS-Cov-2 from saliva. We have paired SARS-CoV-2 detection during isothermal nucleic acid amplification with a mobile, handheld device that detects colorimetric changes during isothermal amplification similar to that of a microplate reader used in previous studies.^12^ This method is capable of detecting purified (synthetic) SARS-CoV-2 RNA template within an order of magnitude of single particle detection (5.25 genomes/reaction) therefore comparable to the theoretical maximum sensitivity of other RT-PCR-based approaches as well as other LAMP-based SARS-CoV-2 detection designs.

COLOR initially received FDA approval for merging LAMP-based testing with a colorimetric endpoint readout that streamlined detection and circumvented the need for highly specialized diagnostic equipment. The majority of FDA-approved LAMP-based assays have only been authorized for samples that have first undergone RNA purification. In order to develop a direct-detection assay from saliva samples that can vary in pH, nucleic acid amplification and pH change must be measured independently. Previous studies have examined the accuracy and sensitivity of endpoint colorimetric changes indirectly by conducting parallel experiments with real-time fluorescent-based assays.^13,33,34^ These approaches, however, cannot directly verify whether a colorimetric readout corresponds to amplification or a spurious pH change. This indirect approach has hindered progress towards developing a direct-detection assay from saliva samples. Our approach, however, provides a platform to independently measure both nucleic acid amplification and colorimetric endpoint readout for every sample and provide a means to determine whether SARS-CoV-2 can be detected directly from saliva.

Using a commercial BioRanger, the time course of the initial luminance derivative peak, L’_MAX1_, corresponded to the time to the threshold of detection for fluorescent-based LAMP assays, while the time course of the second luminance derivative peak, L’_MAX2_, corresponded to the times reported to be required for endpoint color change. These data indicate that our method can detect subtle optical changes at the initiation of LAMP amplification, perhaps due to increased scattering from small particulates of magnesium pyrophosphate, as well as more intense optical changes later in the reaction corresponding to transition of pH through the pK_a_ of the phenol red indicator. The modified BioRanger, therefore, provides an ideal platform for evaluating whether a LAMP-based assay can be implemented for the direct detection of SARS-Cov-2 from saliva.

Direct detection of viral particles from saliva can present a host of additional complications. In addition to fluctuations in pH that can destabilize the viral capsid or cause a spurious color change, saliva contains nucleases that can degrade the viral genome, reducing the amount of virus that can be detected by LAMP. Previous work has shown that encapsulated SARS-CoV-2 is stable in saliva ^20,35,36^ and isothermal nucleic acid amplification efficiency is enhanced when the heat-inactivated saliva matrix is buffered with TE and Tween 20. Processing samples in this way, we implemented a low-cost alternative to VTM that was completely compatible with both luminance detection and endpoint colorimetric detection of isothermal amplification of SARS-Cov-2 nucleic acid.

### Future Implementation

Through the assay that we developed in this study, we were able to directly detect inactivated SARS-CoV-2 from saliva with an LOD of 50 genomes/reaction. This detection sensitivity is similar to a previously-described LAMP-based SARS-CoV-2 direct detection assay used in conjunction with nasopharygeal samples stabilized in VTM.^37^ In October of 2020, COLOR’s rapid LAMP test was deployed at the San Francisco International airport for onsite passenger testing as one of the “Trusted Testing and Travel Partners” in Hawaii’s pre-travel COVID-19 testing program developed to encourage safe travel, tourism and re-opening of the economy. The methods presented here can be adapted in a similar way to provide real-time monitoring as a companion diagnostic to colorimetric endpoint detection of SARS-CoV-2 amplification from easy-to-collect saliva samples. This approach has the potential to reduce the cost and invasive nature of COVID-19 testing and can be broadly applied to point of care detection in field surveillance programs or in remote clinical settings.

## Data Availability

All data used for this research is reported in the manuscript or Supplementary Material.

## Acknowledgements

We would like to thank Dr. John M. Berestecky (Professor, Microbiology and Biotechnology, Kapiolani Community College, Honolulu, HI; Director of the Monoclonal Antibody Service Facility and Training Program) and Dr. Rebecca Kanenaka, (State Certified Microbiologist and CLIA Certified Lab Director, Kapiolani Community College Honolulu, HI) for their roles as project advisors, community outreach and assistance with IBC protocol certification and experimental design, as well as Dr. Reinhold Penner and Dr. Andrea Fleig for reagents and equipment. This project could not have been completed without the generous donation of colorimetric assay reagents from Nathan Tanner (New England Biolabs) who early on during the pandemic also shared LAMP primer sequences used in this study. This research is part of a global initiative, referred to as ‘gLAMP’, to validate LAMP diagnostics for the detection SARS-CoV-2 and we thank all participating members who helped guide our testing protocols from labs across the world. Finally, we express our gratitude to Diagenetix Inc. (Honolulu, HI) for use and deconstruction of commercial BioRanger units and for continued technical support.

## Notes

### Competing Interest Statement

The authors have declared no competing interest.

### Clinical Trial

no clinical trials were conducted

### Author Declarations

IRB approval was waived by the Institutional Review Board - Human Studies Program of the University of Hawaii at Manoa. Based on the research study design, specifically where the samples used did not include identifiers/codes and researchers did not re-identify the samples/data, it was determined that this is Not Human Subjects Research and therefore does not require IRB approval.

