## Supplemental Information for "Real-time optical analysis of a colorimetric LAMP assay for SARS-CoV-2 in saliva with a handheld instrument improves accuracy compared to endpoint assessment"

### Supplementary Information

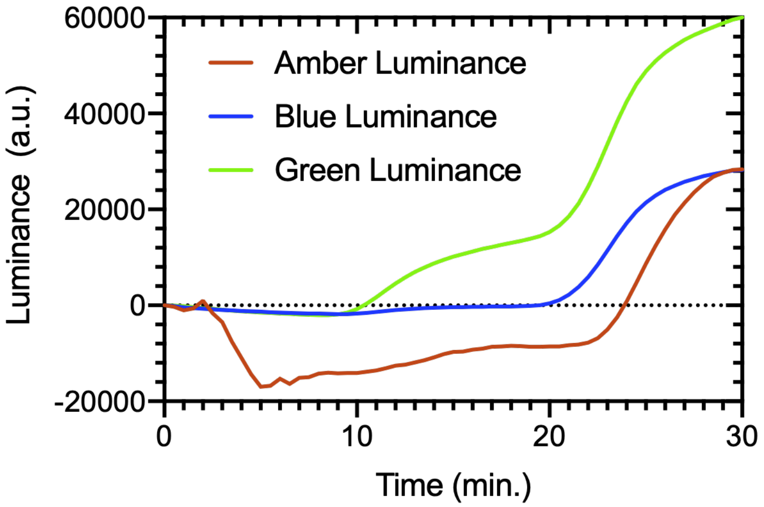


**Supplementary Figure 0. Evaluation of amber, blue and green LEDs for sample illumination and monitoring reactions.** Each curve represents the average luminance values recorded by illumination at 470 nm (blue; n=7), 525 nm (green; n=8) and 590 nm (amber; n=4) channels. All reactions contained NEB's Warmstart colorimetric master mix, a 1:1 quantity of both E1 and N2 primer sets and 1.05 x $10^4$ genome equivalents/reaction of synthetic RNA suspended in DNase/RNase free water. LAMP reactions were performed at 65°C.

**Supplementary Table 1. LAMP primer sequences.** The primers and gene targets are identical those used in New England Biolabs SARS-Cov-2 Rapid Colorimetric LAMP Detection Assay (NEB #E2109) released as a research use only kit in July of 2020.[1]

| Gene target | Primer set | Primer | Nucleotide sequence (5′ → 3′) |
|---|---|---|---|
| SARS-CoV-2 nucleocapsid (N) gene | N2 | F3 | ACCAGGAACTAATCAGACAAG |
| | | B3 | GACTTGATCTTTGAAATTTGGATCT |
| | | FIP | TTCCGAAGAACGCTGAAGCGGAACTGATTACAAACATTGGCC |
| | | BIP | CGCATTGGCATGGAAGTCACAATTTGATGGCACCTGTGTA |
| | | LF | GGGGGCAAATTGTGCAATTTG |
| | | LB | CTTCGGGAACGTGGTTGACC |
| SARS-CoV-2 envelops (E) gene | E1 | F3 | TGAGTACGAACTTATGTACTCAT |
| | | B3 | TTCAGATTTTTAACACGAGAGT |
| | | FIP | ACCACGAAAGCAAGAAAAAGAAGTTCGTTTCGGAAGAGACAG |
| | | BIP | TTGCTAGTTACACTAGCCATCCTTAGGTTTTACAAGACTCACGT |
| | | LF | GCGCTTCGATTGTGTGCGT |
| | | LB | CGCTATTAACTATTAACG |
| Human actin RNA | ACTB | F3 | AGTACCCCATCGAGCACG |
| | | B3 | AGCCTGGATAGCAACGTACA |
| | | FIP | GAGCCACACGCAGCTCATTGTATCACCAACTGGGACGACA |
| | | BIP | CTGAACCCCAAGGCCAACCGGCTGGGGTGTTGAAGGTC |
| | | LF | TGTGGTGCCAGATTTTCTCCA |
| | | LB | CGAGAAGATGACCCAGATCATGT |

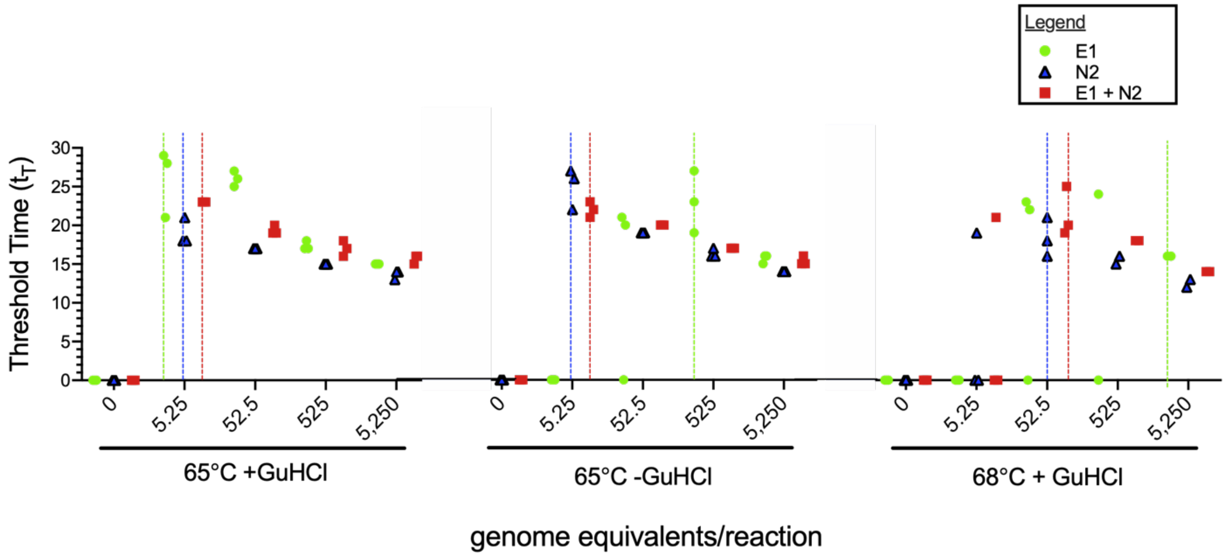


**Supplementary Figure 1. Reaction conditions optimization for detection of synthetic SARS-CoV-2 RNA in spiked water controls by fluorescence-based LAMP**. Three assay replicates (symbols) were performed for each tested concentration (0 – 5,250 ge/reaction). Reactions that failed to amplify are indicated by a symbol at a threshold time of zero ($t_T = 0$). Limit of detection (indicated by dotted line for each primer set) was recorded as the lowest value where SARS-CoV-2 was positively detected in 3/3 reactions.

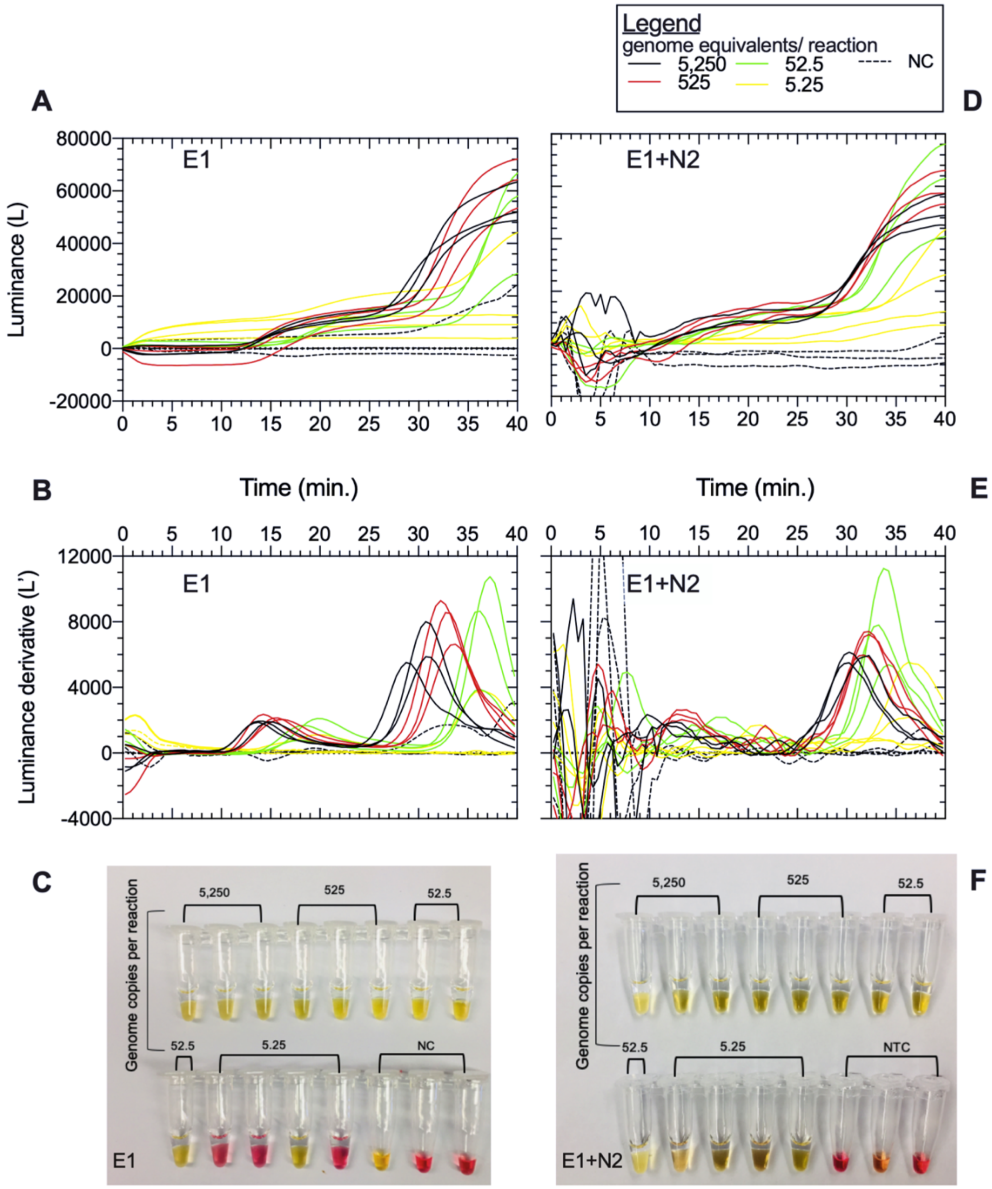


**Supplementary Figure 2. Sensitivity of detecting synthetic SARS-CoV-2 in water and corresponding luminance derivative (L') of single and duplexed primer sets.** The luminance values of LAMP reactions containing E1 (A) and corresponding derivative curves (B) were recorded over the course of 40 minutes for colorimetric LAMP positive controls testing the limit of detection of the tested range (5.25 – 5,250 genome equivalents/ reaction) of synthetic SARS-CoV-2 viral RNA in a 5 µL sample (C). This was repeated identically to evaluate E1 and N2 combined in a LAMP assay (D-F). While duplexed primer sets (E1+N2) were more sensitive than single-plexed (E1) reactions by an order of magnitude (F), the resulting luminance curve (D) and derivative (E) were significantly more noisy with sharp jumps and high amplitude peaks in luminance values especially in the first half of the reaction when a first signature derivative peak is typically distinguishable.

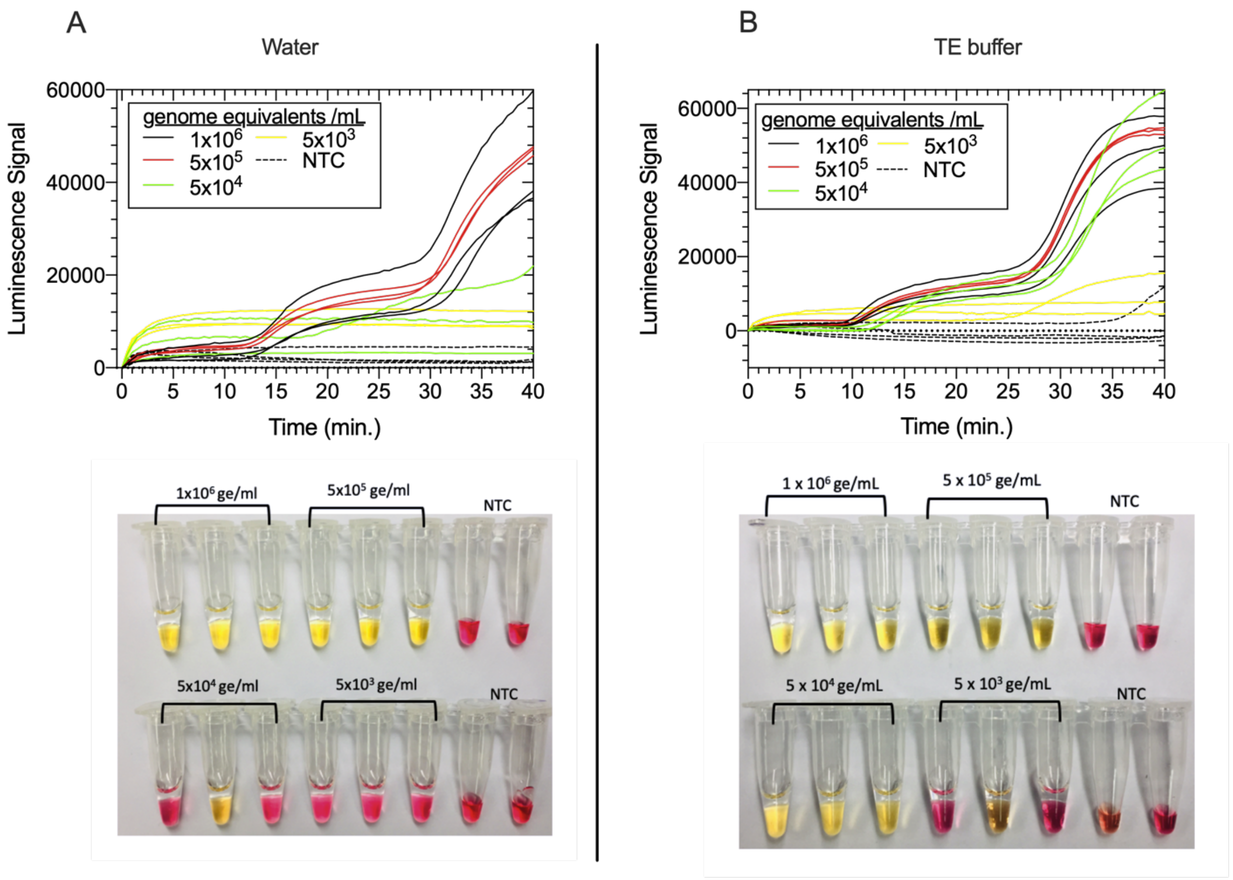


**Supplementary Figure 3. Limit of detection (LOD) of gamma irradiated SARS-CoV-2 in water and TE buffer.**

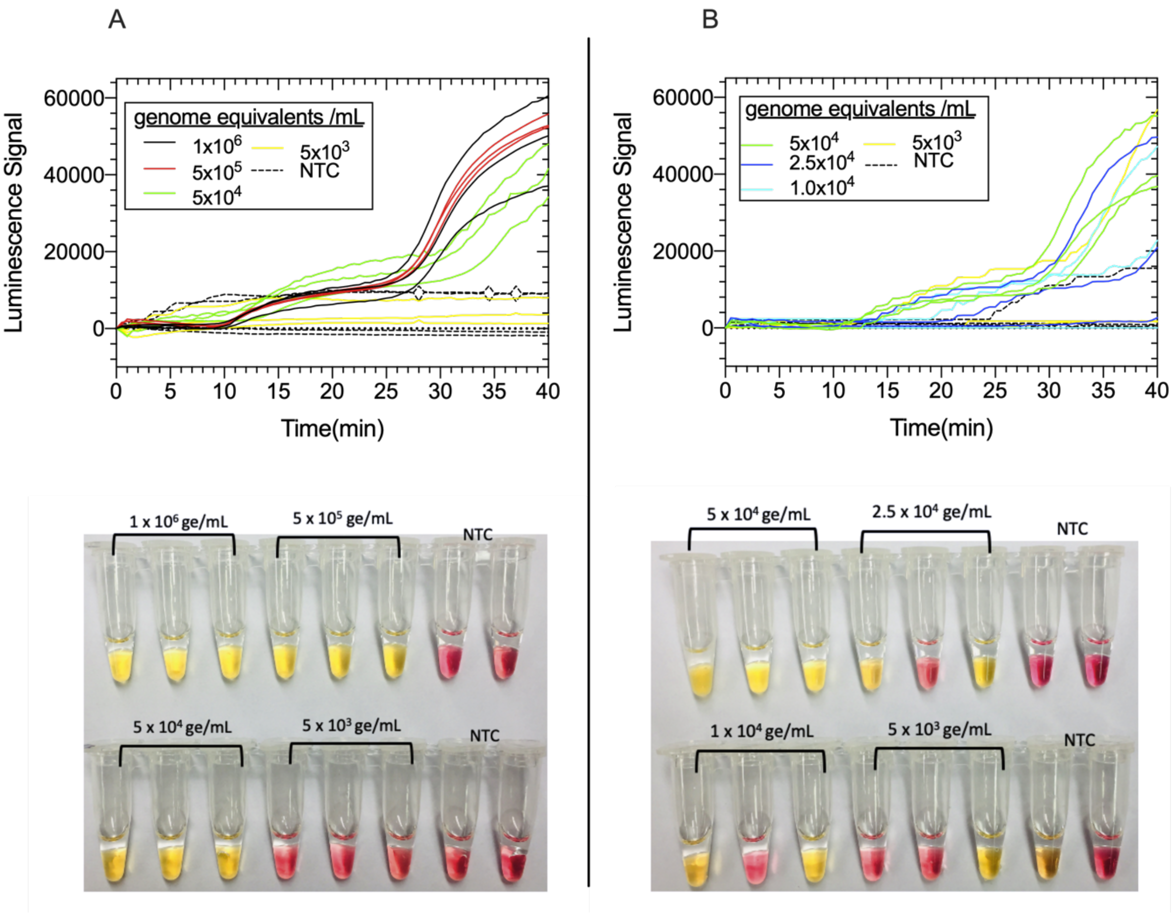


**Supplementary Figure 4. Real-time colorimetric loop-mediated isothermal amplification (LAMP) results for the initial detection limit of gamma-irradiated SARS-CoV-2 N-gene in saliva.** The LOD was initially identified by testing 3 replicates of different concentrations from ~ $10^3$ to $10^6$ genome equivalents/mL (ge/mL) (A) of SARS-CoV-2 in primary saliva samples as 5 x $10^4$ ge/mL (50 ge/ reaction) and verified by later testing 2 and 5 fold below the LOD (B). The LOD was determined as 5 x $10^4$ ge/mL (50 ge/reaction), the lowest concentration of SARS-CoV-2 at which > 95% of assays resulted in positive amplification of luminance signal.

**Supplementary Table 2- Clinical evaluation sample pairing key.**

| Paired Contrived Clinical Samples | |
|---|---|
| 2 (NC) & 8 (1X LOD) | 11 (NC) & 1(2X LOD) |
| 3 (NC) & 10 (2X LOD) | 15 (NC) & 20 (1X LOD) |
| 4 (NC) & 5 (1X LOD) | 16 (NC) & 18 (2X LOD) |
| 6 (NC) & 12 (1X LOD) | 17 (NC) & 14(2X LOD) |
| 9 (NC) & 7 (2X LOD) | 19 (NC) & 13 (1X LOD) |

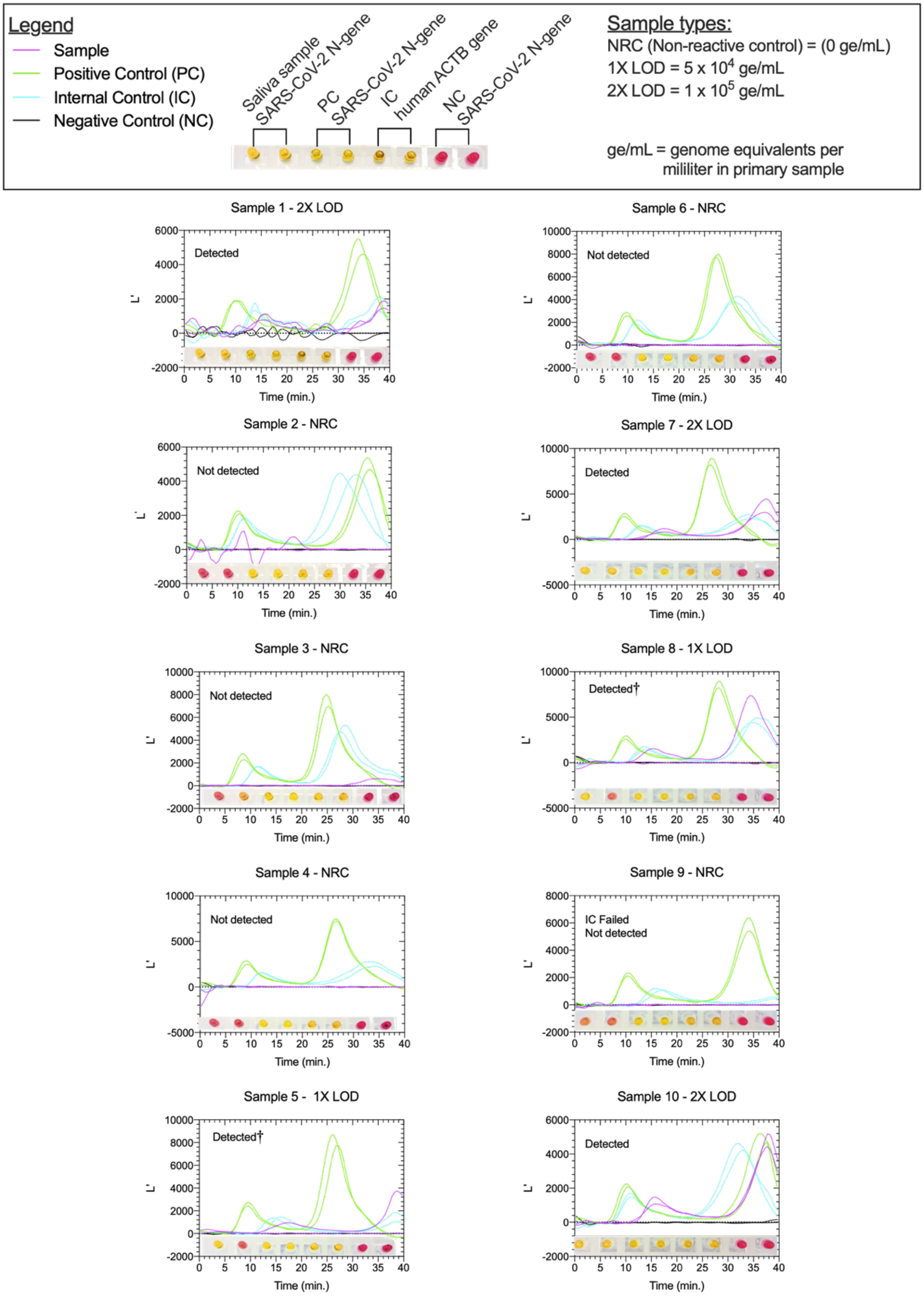
Legend
Sample
Positive Control (PC)
Internal Control (IC)
Negative Control (NC)
Saliva sample SARS-CoV-2 N-gene
PC SARS-CoV-2 N-gene
IC human ACTB gene
NC SARS-CoV-2 N-gene
Sample types:
NRC (Non-reactive control) = (0 ge/mL)
1X LOD = 5 x $10^4$ ge/mL
2X LOD = 1 x $10^5$ ge/mL
ge/mL = genome equivalents per mililiter in primary sample
Sample 1 - 2X LOD
Detected
Sample 6 - NRC
Not detected
Sample 2 - NRC
Not detected
Sample 7 - 2X LOD
Detected
Sample 3 - NRC
Not detected
Sample 8 - 1X LOD
Detected†
Sample 4 - NRC
Not detected
Sample 9 - NRC
IC Failed
Not detected
Sample 5 - 1X LOD
Detected†
Sample 10 - 2X LOD
Detected
L'
Time (min.)

Sample 11 - NRC

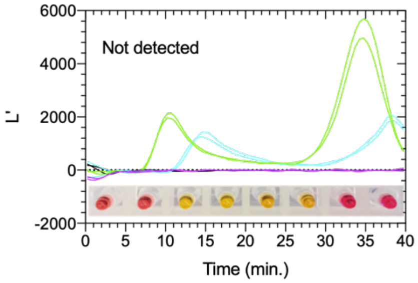


Sample 12 - 1X LOD

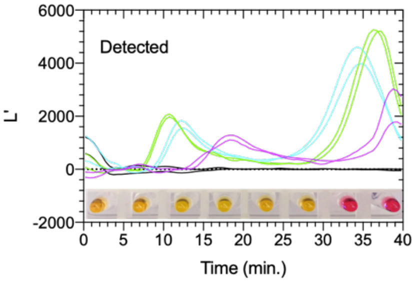


Sample 13 - 1X LOD

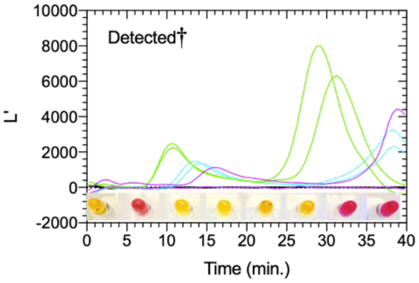


Sample 14 - 2X LOD

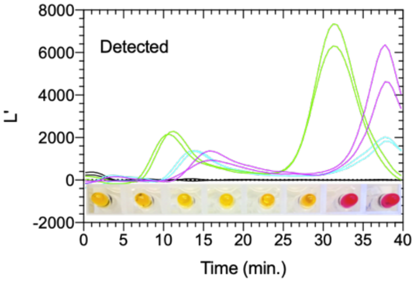


Sample 15 - NRC

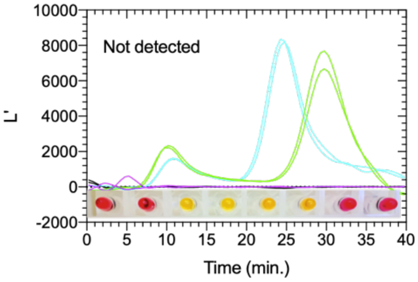


Sample 16 - NRC

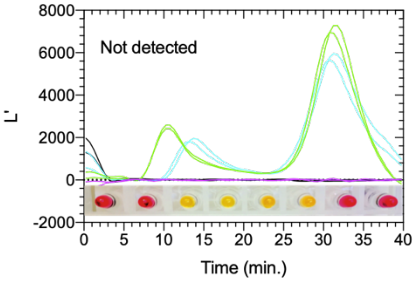


Sample 17 - NRC

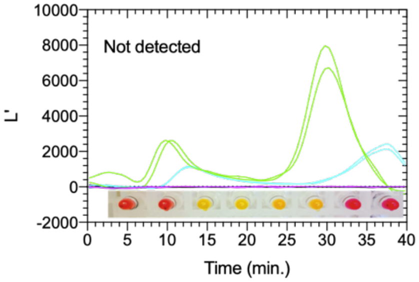


Sample 18 - 2X LOD

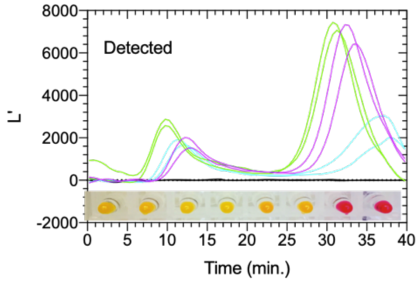


Sample 19 - NRC

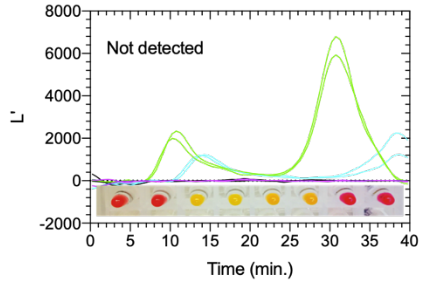


Sample 20 - 1X LOD

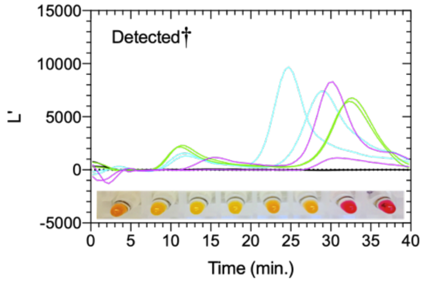

**Supplementary Figure 5. Quantitative and visual results of twenty SARS-COV-2 colorimetric LAMP assay test panels from contrived clinical testing**. A total of 20 test panels (8 assays/ panel), consisting of two replicates each for the tested sample and controls were used to evaluate the performance of our quantitative colorimetric LAMP method to detect SARS-CoV-2 in saliva. A panel is interpreted as 'Detect' if SARS-CoV-2 N-gene is detected in 2/2 replicates, 'Detected†' for 1/2 replicates and 'Non-detect' for 0/2 replicates. Additionally, both replicates must be positive for the internal control and positive control and no detection observed in negative controls.
